# Practice effects persist over two decades of cognitive testing: Implications for longitudinal research

**DOI:** 10.1101/2025.06.16.25329587

**Authors:** Jeremy A. Elman, Erik Buchholz, Rouhui Chen, Mark Sanderson-Cimino, Tyler R. Bell, Nathan Whitsel, Katherine J. Bangen, Alice Cronin-Golomb, Anders M. Dale, Lisa T. Eyler, Nathan A. Gillespie, Eric L. Granholm, Daniel E. Gustavson, Donald J. Hagler, Richard L. Hauger, Diane M. Jacobs, Amy J. Jak, Mark W. Logue, Ruth E. McKenzie, Michael C. Neale, Robert A. Rissman, Rosemary Toomey, Arthur Wingfield, Hong Xian, Christine Fennema-Notestine, Carol E. Franz, Michael J. Lyons, Chandra A. Reynolds, Xin M. Tu, William S. Kremen, Matthew S. Panizzon

## Abstract

**INTRODUCTION:** Repeated cognitive testing can boost scores due to practice effects (PEs). It remains unclear whether PEs persist across multiple follow-ups and long durations. We examined PEs across multiple assessments from midlife to old age in a nonclinical sample.

**METHOD:** Men (N=1,608) in the Vietnam Era Twin Study of Aging (VETSA) underwent neuropsychological assessment across 4 waves from mean age 56 to 74. We leveraged age-matched attrition-replacement (AR) participants to estimate PEs at each wave. We compared cognitive trajectories and prevalence of mild cognitive impairment (MCI) using unadjusted versus PE-adjusted scores.

**RESULTS:** Across follow-ups, a range of 7-12 out of 30 measures demonstrated significant PEs, especially in episodic memory and visuospatial domains. Adjusting for PEs resulted in steeper cognitive decline with up to 29% higher MCI prevalence.

**DISCUSSION:** PEs persist across multiple assessments and decades. The AR-participant method provides accurate sample-specific PE estimates that enable significantly earlier detection of MCI.

## 1. INTRODUCTION

Longitudinal designs are critical for understanding cognitive development and decline [1,2]. In the context of studies on Alzheimer’s disease and Alzheimer’s disease-related dementias (AD/ADRD), the decades-long pathological process and impact of risk factors across the lifespan further underscores the need for long-term longitudinal studies to identify modifiable risk factors, understand variation in disease progression, and evaluate efficacy of treatments [3,4]. However, it has long been acknowledged that performance at follow-up may be artifactually inflated due to practice effects (PEs) [5–7]. Nevertheless, accounting for PEs is not standard practice, which can obscure the true nature of cognitive trajectories and has implications for aging research and AD/ADRD clinical trials [8–10].

PEs are often defined as improved performance at follow-up compared to baseline [11,12]. However, even with observed decline, PEs may still be present when age- or disease-related declines are greater than the magnitude of the PEs [13]. One solution implemented by Rönnlund et al. [14] is to use attrition replacement (AR) participants, new participants recruited from the same population during follow-up study waves who are age-matched to the ongoing longitudinal cohort. This approach is conceptually similar to a randomized controlled trial. By comparing well-matched groups drawn from the same population who differ only on whether they have previously been tested, we can estimate the expected improvement (at the group level) due to practice. Critically, because both groups should experience similar levels of age-related normative decline, we can estimate PEs even when there is an observed *decrease* in scores at follow-up for returnees. This method allows us to create an adjusted follow-up score that can be more appropriately compared to norms or thresholds for impairment, which assume that test scores are unaffected by practice. We have previously shown that using PE-adjusted scores results in earlier detection of mild cognitive impairment (MCI) [13,15,16].

Studies of PEs have typically examined test-retest intervals of less than 5 years, with most being in the range of 6 months to 2 years [17,18]. There has been little investigation of PEs in cohorts that have followed individuals for extended periods of time (i.e., over 10 years, but see [19] and [20] for notable exceptions). Extended longitudinal follow-ups that account for PEs are necessary to clarify cognitive trajectories across critical transition periods such as midlife to old age, especially with increased recognition that disease processes can begin decades prior to clinical onset. The Vietnam Era Twin Study of Aging (VETSA) presents a rare opportunity to examine PEs on cognitive performance in ∼1,600 individuals assessed over 4 study waves. Here, we extend the method [14] that we previously applied to two waves of VETSA data [15] using a generalized approach that more flexibly handles complex testing schedules and missingness patterns. We examined how PEs evolve over two decades and compared cognitive trajectories and MCI prevalence using unadjusted versus PE-adjusted scores.

## 2. METHODS

### 2.1 Participants

Participants were 1,608 individuals tested at one or more of the 4 completed VETSA study waves (**Table 1**). VETSA is an on-going longitudinal study of cognitive and brain aging beginning in middle age [21–23]. Participants were members of the Vietnam Era Twin Registry, a national, community-dwelling sample of male-male twins who served in the U.S. military during the Vietnam era (1965-1975) [24]. All Registry members were invited to participate in the Harvard Drug Study [25], for which ascertainment was not based on any diagnostic or substance use criteria. VETSA participants were then randomly recruited from the Harvard Drug Study sample. At baseline, VETSA participants were similar to American men in their age cohort with respect to health, education, and lifestyle characteristics based on Center for Disease Control and Prevention data [26], and nearly 80% reported no combat exposure during their military service [27].

**Table 1.** Demographic characteristics of the sample.

|  |  |
| --- | --- |
| N Total | 1608 |
| Age (years); mean (SD) |  |
| *Wave 1* | 55.94 (2.43) |
| *Wave 2* | 61.72 (2.45) |
| *Wave 3* | 67.56 (2.53) |
| *Wave 4* | 73.09 (2.09) |
| Education (years); mean (SD) | 13.85 (09) |
| Age 20 AFQT (percentile); mean (SD) | 59.95 (23.11) |
| Race; n (%) |  |
| *American Indian or Alaskan Native* | 5 (0.3%) |
| *Native Hawaiian or Pacific Islander* | 2 (0.1%) |
| *Black or African-American* | 88 (5.5%) |
| *White* | 1492 (92.8%) |
| *More than one race* | 21 (1.3%) |
| *Decline to answer* | 2 (0.1%) |
| Ethnicity; n (%) |  |
| *Hispanic* | 44 (2.7%) |
| *Non-Hispanic* | 1564 (97.3%) |
| Testing interval (years); mean (SD) |  |
| *Wave 1 to 2* | 5.69 (0.69) |
| *Wave 2 to 3* | 5.91 (0.76) |
| *Wave 3 to 4* | 5.63 (0.60) |

Note: AFQT = Armed Forces Qualifying Test

Of the 1,608 individuals in the current study, 1,291 were enrolled and tested during the wave 1 baseline assessment. At waves 2 and 3, attrition replacement participants age-matched to the ongoing sample were recruited from the VET Registry and tested for the first time (wave 2 n=193; wave 3 n=124). These participants were then invited for follow-up at all subsequent waves (see **Supplemental Figure S1A** for all patterns of assessments). On average, participants were 56 years of age (range 51-61) at wave 1, 62 years (range 56-67) at wave 2, 68 years (range 61-73) at wave 3, and 74 (range 67-79) at wave 4 (see **Supplemental Figure S1B** for age distributions by wave). The average time between wave 1 and 2 was 5.7 years, with 5.9 years between waves 2 and 3, and 5.6 years between waves 3 and 4.

The study was performed in accordance with the ethical standards per the 1964 Declaration of Helsinki and later amendments. Informed consent was obtained from all participants and institutional review boards at both sites approved all study procedures.

### 2.2 Cognitive tests and measures

Cognitive measures included in the current analysis were a set of 30 component scores from neuropsychological tests covering multiple cognitive domains (see **Table 2** and **Supplemental Material** for full list of tests and measures). For consistency, scores from tests where lower values indicate better performance (e.g., reaction time) were reverse-coded so that higher scores uniformly indicate better performance. The same tests were assessed at all waves with 2 exceptions: spatial span (waves 1, 2, and 3 only) and Boston Naming Test (waves 3 and 4 only). Therefore, all 30 measures have 1 follow-up, with 29 and 28 measures having two and three follow-ups, respectively. Additionally, young adult general cognitive ability (GCA) was assessed with the validated Armed Forces Qualification Test (AFQT) [28], which is highly correlated with standard IQ scales (r=.84) [29]. AFQT was administered during military induction at average age 20 (hereafter referred to as “age 20 GCA”).

**Table 2.**
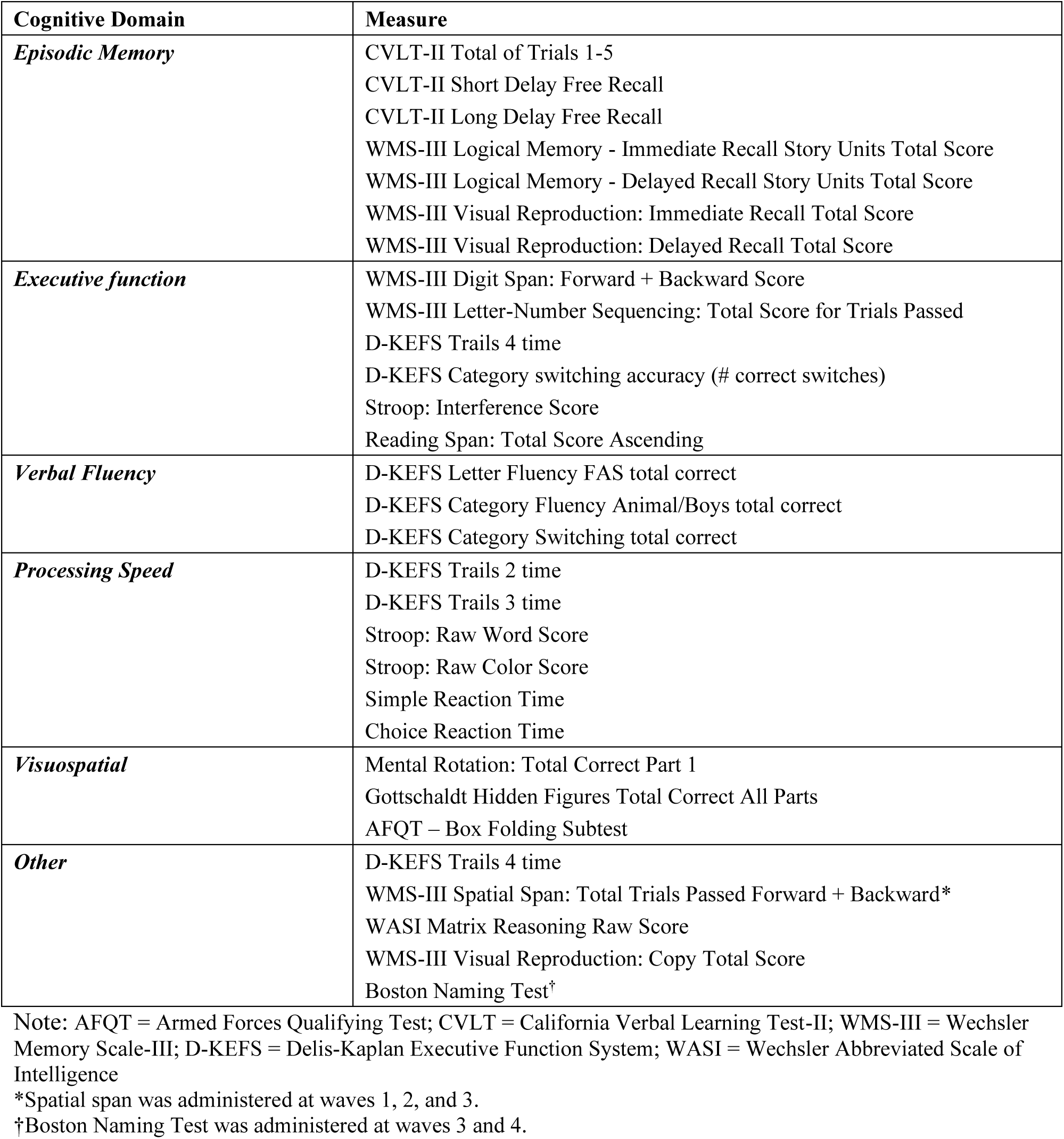
Neuropsychological test scores included in practice effects estimation. Individual scores are grouped based on the cognitive composite that they contribute to. Measures listed in the “Other” category were not used to calculate cognitive factor scores but were used to classify mild cognitive impairment.

### 2.3 Estimation of practice effects

PEs were estimated for each of the 30 measures using generalized estimating equations (GEE). We opted for this class of semiparametric regression models because it requires no assumptions about data distributions, such as normality, to provide valid inferences for virtually all data distributions arising in practice [30].

Let 𝑌_𝑖𝑎𝑤_ denote the score on a cognitive measure at the *a*^th^ assessment of subject *i* occurring at wave *w*:

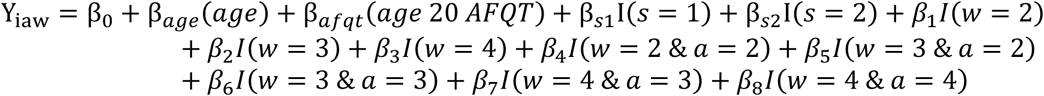

Here, 𝐼(⋅) denotes an indicator function that takes the value 1 if the condition inside is true, and 0 otherwise. Wave *w* and Assessment *a* can take values from 1 to 4. Importantly, the inclusion of AR participants means there are groups of individuals at each follow-up wave that differ in number of prior assessments. Therefore, additional indicator variables capture the interaction between Wave and Assessment (with the necessary condition that *a≤w*). Participants may miss a study wave but then return for follow-up at subsequent waves. To account for the potential impact of longer intervals between assessments on PEs, we include the Skip variable *s* that can take the values 1 or 2 to indicate whether they missed 1 or 2 waves immediately prior to the current assessment (only missed assessments after enrollment are counted).

We additionally adjusted for age and age 20 GCA. Adjusting for age allows us to estimate PEs independent of normative age-related decline. Previously, we found that the attrition replacement group at Wave 2 had significantly lower young adult (age 20) GCA scores than the group enrolled at Wave 1, which could result in artificially inflated PEs. Given that participants were randomly recruited from the same population (i.e., the VET Registry), this is likely due to random sampling variation. However, this adjustment helps account for potential long-standing differences in performance. Details on interpretation of each model coefficient are provided in **Supplemental Methods and Supplemental Table S1**.

### 2.4 Adjusting test scores for practice effects

For a given individual, we can calculate the expected PE for a measure by taking the coefficients corresponding to the relevant wave and assessment number, and whether they have skipped any previous assessments. The resulting “adjustment value” is then subtracted from their observed score. This results in a score that we would expect if they had been taking the test for the first time. As an example, consider two individuals at Wave 4. The first has completed all four waves of testing, so we can adjust their score on a given measure at Wave 4 by subtracting the value of β_8_ (i.e., the PE for someone completing a 4^th^ assessment at Wave 4) from their observed score. The other individual missed Wave 3, so it would be their third assessment. For this person, we would instead use β_7_ and add to that the estimated effect of skipping one prior assessment (i.e., β_s1_). See **Supplemental Methods** for further discussion of selecting coefficients for practice effect adjustment.

### 2.5 Cognitive factor score composites

We have previously shown that composites such as cognitive factor scores can improve reliability and prediction of cognitive decline [31]. Therefore, we were interested in understanding how adjusting individual scores could impact cognitive composites. We calculated composites for 5 cognitive abilities from both adjusted and unadjusted scores: episodic memory, executive function, verbal fluency, processing speed, and visuospatial ability. Details of the factor models from which these composites were derived are described in previous publications [31–34] and in **Supplemental Methods**. Higher values reflect better performance in each domain. We used paired t-tests to compare cognitive composites calculated from adjusted and unadjusted scores at each wave.

### 2.6 Classification of mild cognitive impairment

Classification of MCI was compared both before and after adjusting for PEs. We defined MCI according to the Jak/Bondi approach as described previously [35,36] and in **Supplemental Methods**. To ensure that MCI classification captured cognitive decline rather than lifelong low ability, neuropsychological scores were adjusted using early adult GCA (age 20 AFQT) as a covariate. Impairment was defined as having 2+ measures within a domain >1.5 SD below age-based normative means. Individuals with an impaired memory domain were classified as amnestic MCI (aMCI), and those with impairments in domains other than memory were classified as non-amnestic MCI (naMCI). Differences in MCI classification based on adjusted or unadjusted scores were assessed at each wave with McNemar’s χ^2^ test with 1 degree of freedom.

## 3. RESULTS

### 3.1 Practice effects on cognitive tests across 4 study waves

Models included all data and patterns of assessments, but here we focus on reporting estimates for individuals who attended all 4 waves. This was the most common pattern of participation and allowed us to examine the evolution of PEs over the longest follow-up period and greatest number of assessments. **Figure 1** presents PE estimates and confidence intervals for each of the 3 follow-up assessments for those that completed all waves. The PE estimates can be interpreted as the expected boost in performance an individual receives from having taken the test a given number of times previously compared to someone of a similar age and young adult GCA taking the test for the first time. At the first follow-up, 12 of 30 measures demonstrated significant PEs ranging in magnitude from 0.14 SD units to 0.29 SD units. At the second follow-up, 7 of 29 measures demonstrated significant PEs ranging from 0.16 SD units to 0.34 SD units. At the third follow-up, 8 of the 28 measures demonstrated significant PEs ranging from 0.23 SD units to 0.35 SD units. PEs. Although the number of significant PEs was smaller at later follow-ups, this is likely due to smaller sample size at these visits. Visuospatial and episodic memory measures consistently showed the strongest effects, and measures that showed significant PEs at later follow-ups typically, but not always, showed significant PEs at earlier follow-ups.

**Figure 1.**
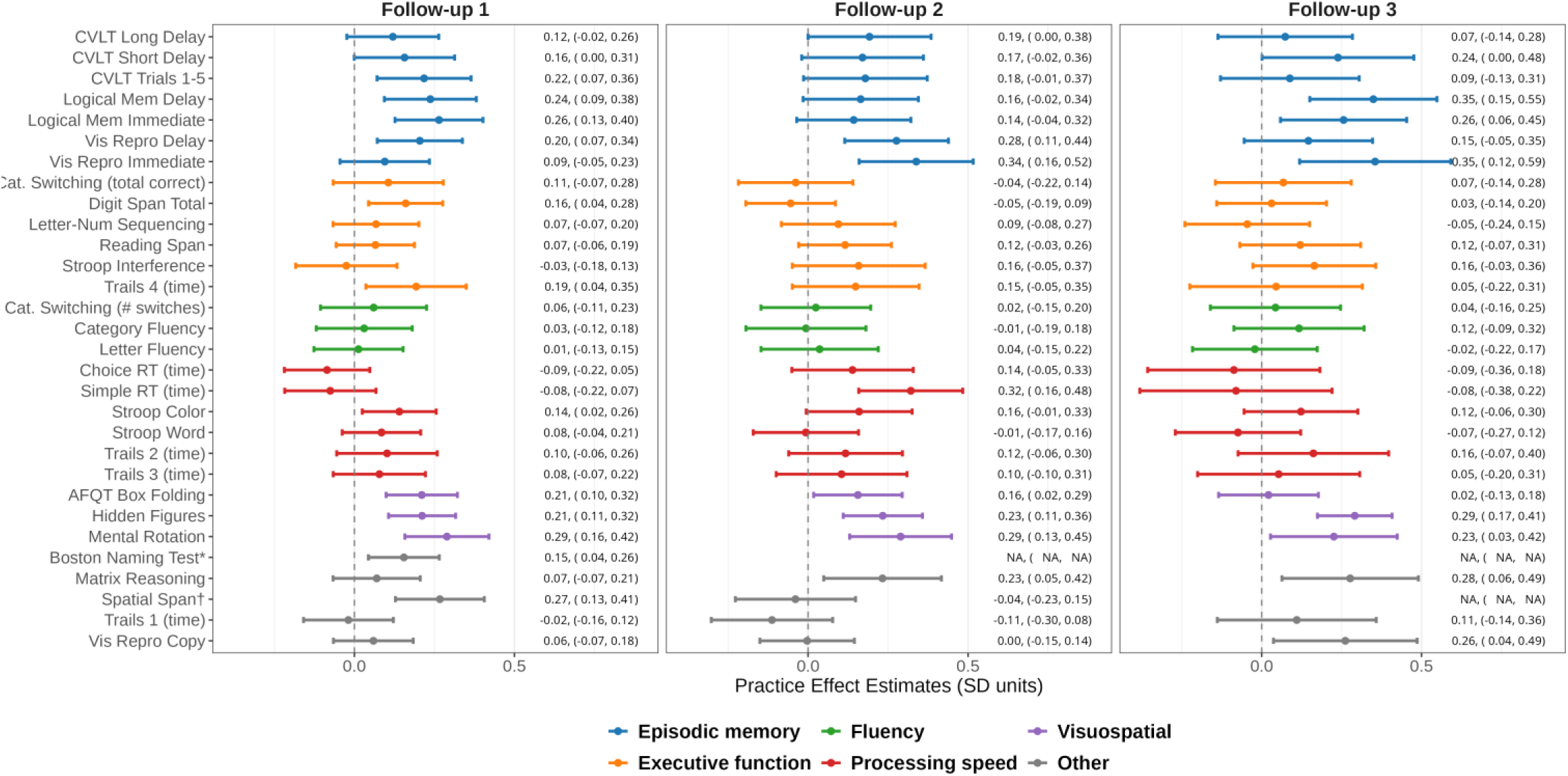
Practice effect estimates across follow-up assessments. The forest plot presents practice effect estimates and 95% confidence intervals for practice effect estimates at each follow-up. The first, second and third follow-ups occurred at waves 2, 3, and 4, respectively, for all tests except the Boston Naming Test. The Boston Naming Test was introduced at wave 3, so the first follow-up occurred at wave 4. Models included all participants and assessment patterns but only estimates corresponding to individuals that participated in all four study waves are presented in this figure (i.e., column 1 = 𝛽_4_, column 2 = 𝛽_6_, column 3 = 𝛽_8_). Coefficients can be interpreted as the expected boost in performance in standard deviation units expected for returnees at a given wave and given number of prior assessments compared to a test-naïve participant of a similar age and young adult general cognitive ability level. All items were coded such that higher values reflect better performance. See Table 2 for full names of measures in each domain. AFQT=Armed Forces Qualifying Test. * Boston Naming Test was administered at waves 3 and 4 only. † Spatial Span was administered at waves 1, 2, and 3 only.

### 3.2 Impact on cognitive factor scores

PE-adjustment led to significantly lower composite scores across all follow-up waves for all domains (all ps < 0.05), indicating that unadjusted scores can overestimate cognitive performance in these domains. Consistent with results from individual test measures, differences in performance were most notable for composites of episodic memory (ranging between -0.21 to -0.25 SD units) and visuospatial ability (ranging between -0.19 to -0.22 SD units), and were weakest in the fluency domain (ranging between -0.04 to -0.07 SD units). See **Figure 2** and **Supplemental Table S3** for full results of comparisons across waves.

**Figure 2.**
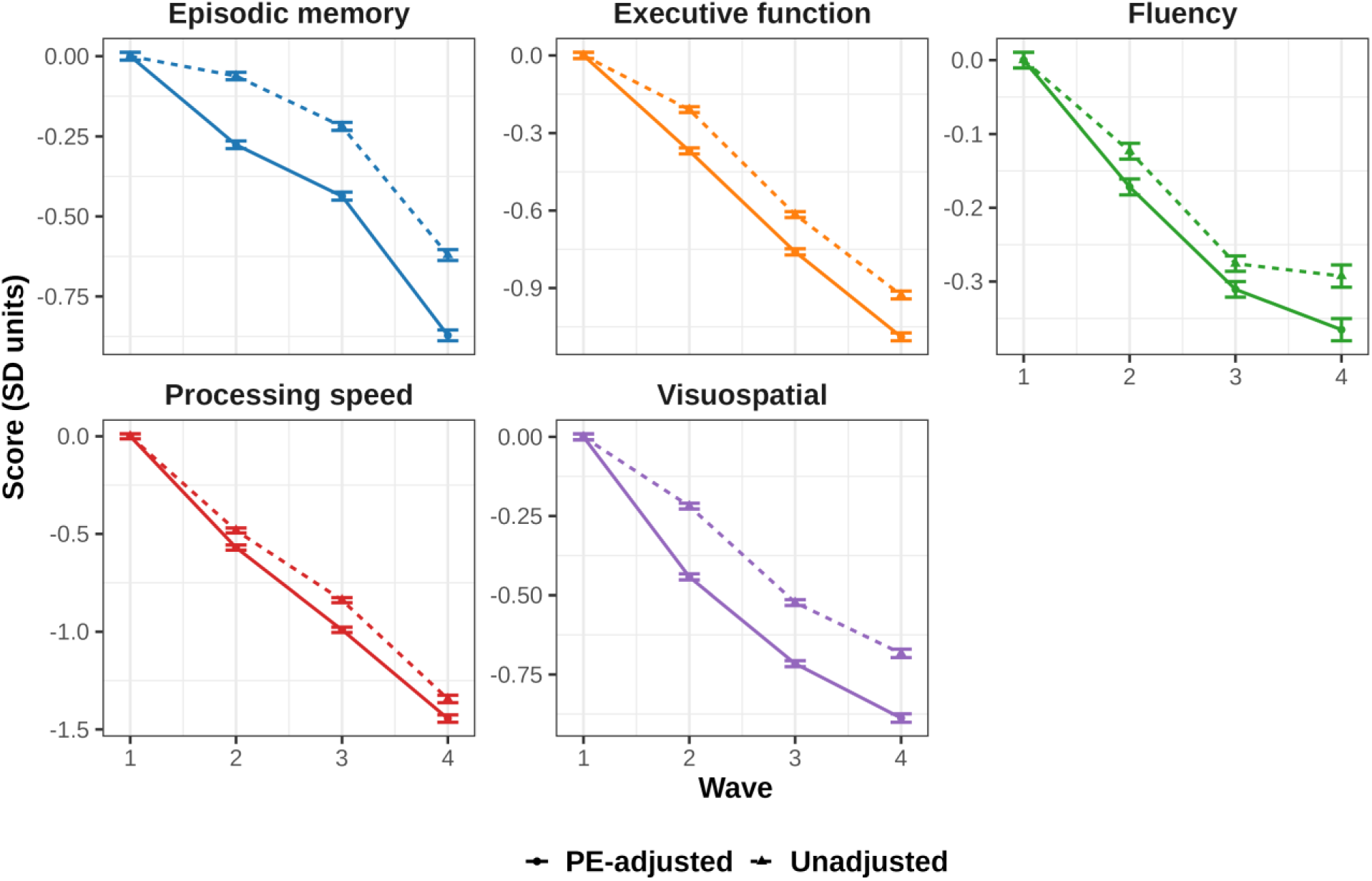
Plots of cognitive factor score trajectories. Means and within-subject standard errors for cognitive factor score composites calculated from unadjusted (triangles and dashed lines) versus practice effect-adjusted (dots and solid lines) scores. Scores were standardized using the sample means and standard deviations at wave 1.

### 3.3 Impact on classification of MCI

We next examined the impact of adjusting for PEs on the rate of MCI at follow-up assessments (**Figure 3**). Adjusting for PEs significantly increased the prevalence of MCI at all follow-up waves (wave 2: 12.3% vs. 15.6%, χ^2^=32.60, p<0.001; wave 3: 15.1% vs. 18.1%, χ^2^=23.67, p<0.001; wave 4: 16.3% vs. 21.0%, χ^2^=33.23, p<0.001). These increases suggest that failure to adjust for PEs masks clinically meaningful cognitive decline, particularly in long-term follow-up. Consistent with findings that measures in the memory domains exhibited the strongest PEs, the increased rates of MCI were largest for amnestic MCI. After PE adjustment, the rate of amnestic MCI was significantly higher at all follow-up waves (wave 2: 8.5% vs 11.4%, χ^2^=26.036, p<0.001; wave 3: 9.6% vs 13.2%, χ^2^=25.289, p<0.001; wave 4: 12.6% vs 16.6%, χ^2^=22.042, p<0.001). Rates of non-amnestic MCI were significantly higher at waves 2 and 4, but not wave 3 (wave 2: 4.6% vs 5.2%, χ^2^=5.786, p=0.016; wave 3: 6.6% vs 6.5%, χ^2^=0.125, p=0.724; wave 4: 4.9% vs 6.3%, χ^2^=9.600, p=0.002).

**Figure 3.**
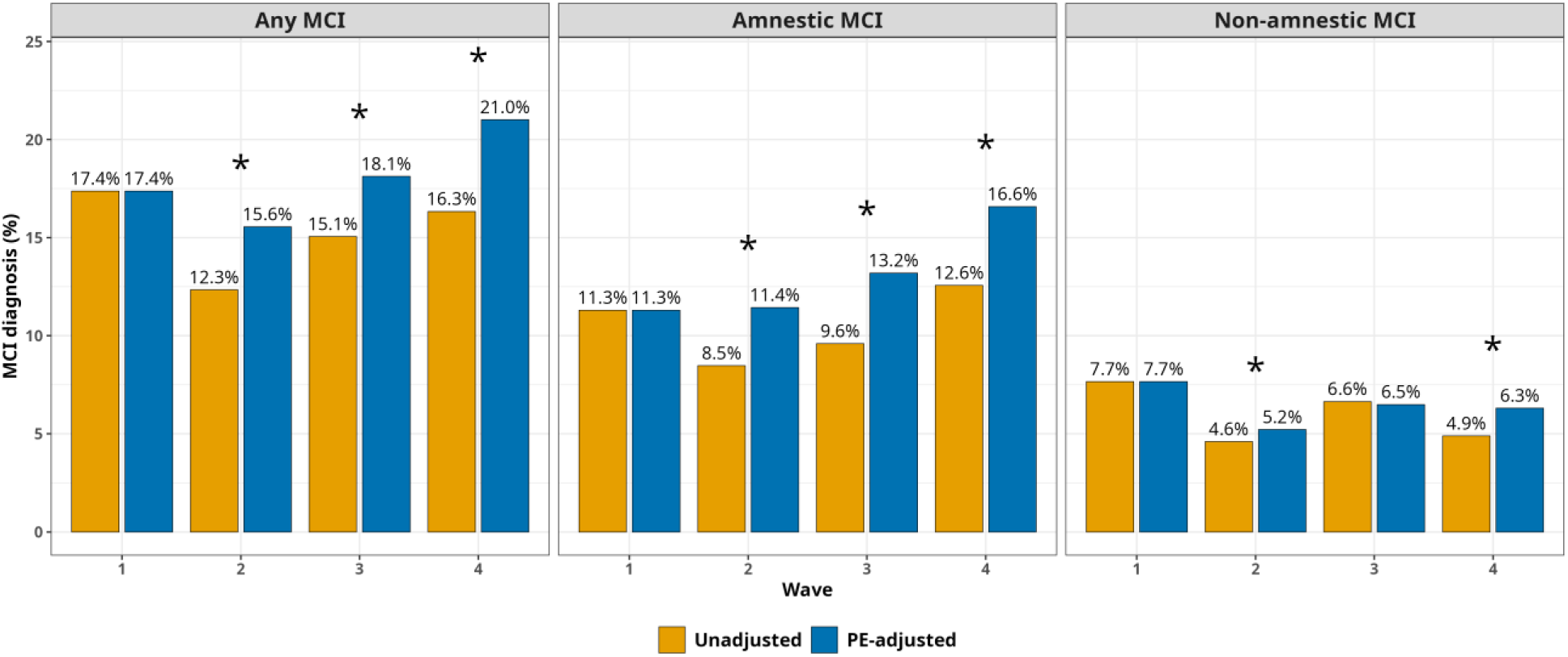
Rates of mild cognitive impairment. Bar plots present prevalence of any mild cognitive impairment (left panel), amnestic MCI (middle panel) and non-amnestic MCI (right panel) at each wave based on unadjusted (orange) and practice effect-adjusted (blue) test scores. Asterisks indicate significant (p<0.05) differences in prevalence between unadjusted and adjusted rates within a wave.

## 4. DISCUSSION

These results demonstrate the persistence of PEs on multiple cognitive measures over extended periods of time and multiple assessments. Our findings are broadly consistent with other studies in that PEs were most significant at first follow-up with fewer significant PEs at later follow-ups [12,19,37–40]. However, this was not due to a consistent decrease in effects sizes and may be explained by smaller sample sizes at these waves, thus PEs at later follow-ups should not be discounted. Importantly, even when PEs were modest and non-significant, we found that they could have meaningful impacts on downstream analyses. Adjusting for Pes revealed significantly greater decline in cognitive trajectories as well as increased rates—and hence, earlier detection—of MCI at all waves. Here, we address 5 issues that are relevant to the present findings and are frequently raised in regard to PEs.

### First, which domains are most susceptible to PEs and how does the magnitude of PEs compare with other studies?

Consistent with prior studies, we found PEs were most apparent and most persistent in the memory domain, whereas fluency, executive function, and speed showed the smaller effects [11,12,17,37,41]. We additionally found strong PEs in the visuospatial domain, which is consistent with some studies [12] but not others [17,37]. However, direct comparisons across studies may not be entirely appropriate. PEs vary depending on the specific tests used, length of retest interval, number of follow-ups, and participant characteristics such as general cognitive ability, age, sex, race/ethnicity, diagnostic status, and presence of pathology [11,37,42,43]. Underscoring this point, we found that the magnitude of PEs differed across individual measures within the same domain. Because estimates from one study may not apply to another, it is ideal to estimate study-specific PEs. Estimating PEs using an AR-participant approach builds in a mechanism for deriving valid estimates that are specific to the sample characteristics (e.g., age and other demographics) and study design (e.g., test battery, retest interval).

### Second, how long do practice effects last?

Our results provide evidence of PEs across testing intervals of at least 5-6 years. Although this testing interval is comparatively long, other studies have found evidence of PEs after 7 years [14,17,19,44]. Regarding how PEs persist across number of follow-ups, our study provides evidence that PEs continued to influence performance through Wave 4. One of the few studies examining PEs for multiple assessments over a comparable period of time also found PEs on an intelligence test up to the fourth wave [19,40]. It should be noted that most prior studies finding a lack of PEs at follow-up did not use AR-participant approaches, which may underestimate or fail to detect PEs (i.e., they may be obscured by greater decline at later timepoints). Moreover, performance on certain tests may have plateaued at later follow-ups as participant performance reached ceiling on the administered tests, preventing any further practice-related boosts [12,37–39].

### Third, what are the mechanisms underlying PEs?

There are likely to be multiple mechanisms, with varying contributions depending on the test. Participants may explicitly remember certain content from prior assessments (e.g., aspects of the story given during Logical Memory), which aids their performance. Yet on some tests such as Digit Span, it is highly unlikely participants are remembering specific sequences of digits. In these scenarios, PEs may be driven by greater familiarity with the *context.* This could include reduced test anxiety due to familiarity with the testing environment, procedural memory for tasks with a motor component, or identifying effective test-taking strategies. These context-related factors may explain why even individuals with severe episodic memory impairment can sometimes still benefit from practice [45]. It is worth noting, however, that the AR-based approach used here to estimate and adjust for PEs here does not hinge on knowing their underlying causes, but quantifies their aggregate effect.

### Fourth, how do PEs affect detection of MCI?

We found that adjusting for PEs resulted in significant increases in the rate of MCI at all waves, driven primarily by increases in amnestic MCI. This supports our prior findings that not accounting for PEs underestimates or delays diagnosis of MCI [13,15,16,46]. As shown elsewhere, accounting for PEs to identify MCI earlier can reduce the required sample size of clinical trials that use progression to impairment as an endpoint, resulting in multi-million dollar cost savings and allowing for more accurate estimate of treatment effects [8,9,16]. Of course, adjusting scores downward necessarily means that more people will be below the impairment threshold, but in an independent sample we showed that PE-adjusted diagnoses resulted in lower rates of reversion and increased concordance with biomarker-positivity [16,46], providing evidence that the increased prevalence of MCI was not driven by false positive diagnoses. Finally, earlier detection is critical to enabling early intervention, which is likely to improve treatment effectiveness.

### Fifth, how does the AR-participant approach compare with other PE methods?

We utilized an AR-based approach to estimate group-level PEs representing the average increase in score expected from having taken a given test once or several previous times. Our study extends prior AR-based methods [14-16,46,47] to better handle larger numbers of follow-up assessments and patterns of missingness. Some approaches employ multiple tests within short time frames (e.g., 1 week test-retest interval) [18]. Although bearing a similar name, these short-term PE studies have a different purpose than our approach, namely, for prognosis or predicting future decline. Other methods, such as the Reliable Change Index (RCI) [48] or Standardized Regression Based (SRB) change indices [49], focus on analysis of *change scores*. In contrast, our approach focuses on obtaining *adjusted follow-up scores* that, when compared to norm-based thresholds, enable earlier identification of impairment. Despite the shared use of the term “practice effects,” these approaches are therefore not comparable nor are they in competition as they have different goals. Importantly, the AR-based approach can disentangle PEs from normative decline, which avoids underestimation of PEs. Moreover, the estimates are study-and sample-specific, which enable earlier and more accurate identification of MCI.

We note some limitations of the current study. The VETSA is an all-male sample and primarily non-Hispanic White, limiting its generalizability to other samples. Similar results have been found over 1- to 5-year intervals in mixed-sex samples [14,16,46], but as noted, the AR approach is meant to be applied within individual studies to obtain study-specific PEs. The AR-approach does add costs and time to complete a study, yet a relatively small number of replacement participants (∼10% of the sample) was sufficient to estimate PEs specific to our study. Importantly, we have previously described an approach to obtain “pseudo-replacements” [16] that can also be used in studies when dedicated AR participants were not part of an initial study design. The fact that the method determines PEs at the group, rather than the individual level may also be a limitation. However, as described above, approaches that estimate individual-level PEs typically have a different goal (i.e., prognosis or prediction of decline). It is not clear that there is an approach to obtain individual-level PEs for the purposes of adjusting follow-up scores that can properly disentangle differential practice from differential decline.

In summary, with the AR-participant approach we were able to demonstrate that PEs arising from repeated cognitive testing may persist across multiple assessments and over periods as long as 20 years, with substantial impacts on cognitive trajectories and detection of MCI. This approach uniquely allows for detection of PEs even when observed scores decline and extends our prior work to handle multiple follow-ups and patterns of missingness. Adjusting follow-up scores enables earlier detection of progression to MCI, which has the potential for substantial cost savings in clinical trials [10,16] and allows for earlier and perhaps more effective intervention. These results underscore the importance of accounting for PEs in longitudinal studies of aging and clinical trials aimed at slowing cognitive decline.

## ACKNOWLEDGMENTS

This work was supported by the National Institute on Aging at the National Institutes of Health (grant numbers R01s AG076838, AG022381, AG050595, AG064955; and K01 AG063805). The content of this manuscript is the responsibility of the authors and does not represent official views of NIA/NIH, or the Veterans’ Administration. Numerous organizations provided invaluable assistance in the conduct of the VET Registry, including: U.S. Department of Veterans Affairs, Department of Defense; National Personnel Records Center, National Archives and Records Administration; Internal Revenue Service; National Opinion Research Center; National Research Council, National Academy of Sciences; the Institute for Survey Research, Temple University. The authors gratefully acknowledge the continued cooperation of the twins and the efforts of many staff members.

## CONSENT STATEMENT

Informed consent was obtained from all participants and institutional review boards at the University of California San Diego and Boston University approved all study procedures. data; or in the writing of the report.

## DATA AVAILABILITY

Instructions for data access requests are available on the VETSA website (https://psychiatry.ucsd.edu/research/programs-centers/vetsa/researchers.html). Access to data from military induction can be requested from the Vietnam Era Twin Registry (https://www.seattle.eric.research.va.gov/VETR/Investigator_Access.asp).

