## Supplemental Material for "Practice effects persist over two decades of cognitive testing: Implications for longitudinal research"

### SUPPLEMENTAL METHODS

#### *Interpreting coefficients from the practice effects model*

For convenience, we provide the equation used to estimate practice effects here. Let $Y_{iaw}$ denote the score on a cognitive test measure at the $a^{th}$ assessment of subject $i$ occurring at wave $w$:

$$\begin{aligned} \mathrm{Y_{iaw}} = {} & \beta_0 + \beta_{age}(age) + \beta_{afqt}(age\ 20\ AFQT) + \beta_{s1}\mathrm{I}(s = 1) + \beta_{s2}\mathrm{I}(s = 2) + \beta_1 I(w = 2) \\ & + \beta_2 I(w = 3) + \beta_3 I(w = 4) + \beta_4 I(w = 2\ \&\ a = 2) + \beta_5 I(w = 3\ \&\ a = 2) \\ & + \beta_6 I(w = 3\ \&\ a = 3) + \beta_7 I(w = 4\ \&\ a = 3) + \beta_8 I(w = 4\ \&\ a = 4) \end{aligned}$$

The coefficients from the GEE models described above can be interpreted as the expected difference in score on a particular measure associated with the relevant wave, assessment number, and whether any prior waves were skipped (see **Supplemental Table S1** for interpretation of model terms). With the inclusion of terms for age and age 20 AFQT, the descriptions of coefficients below should be interpreted as being conditional on, or controlling for, age and young adult general cognitive ability. Unless otherwise noted, coefficients are interpreted relative a first assessment at wave 1 with no skipped waves, conditional on age and age-20 AFQT.

The coefficients $\beta_1$ and $\beta_2$ capture the main effects of waves 2 and 3. However, due to the inclusion of interaction terms for number of assessments at each wave, these more precisely correspond to the reference level of assessment at waves 2 and 3. In this case of both waves 2 and 3, the reference assessment is the first time taking the test ($a = 1$). Therefore, these coefficients can be interpreted as the effect of taking the first assessment at wave 2 or 3 for the attrition replacement samples at the respective waves. More concretely, this is the difference in baseline scores of attrition replacements at waves 2 and 3 compared to the baseline score of the original sample, conditional on age and age-20 AFQT. Similarly, the intercept $\beta_0$ reflects the mean score of individuals whose first assessment was at wave 1 (i.e., baseline for the original sample).

Of greatest interest are coefficients $\beta_3$ through $\beta_8$, which can be interpreted as practice effect estimates: the expected boost in performance gained from having taken a test a given number of times prior to a given wave compared to someone of a similar age and age-20 AFQT taking the test for the first time. The interpretation of $\beta_3$ is somewhat different than $\beta_1$ and $\beta_2$ despite not having an indicator for Assessment $a > 1$. All three coefficients are coded as main effects of their respective waves. However, because no replacement participants were recruited at wave 4, adding a separate indicator for ($w = 4\ \&\ a = 2$) would induce collinearity (i.e., values for this term would be identical to the main effect term for $w = 4$). Therefore, the reference level corresponds to a second assessment at wave 4. Therefore, this coefficient can be interpreted as the PE for individuals who are taking their second assessment at wave 4 compared to an individual taking the test for the first time, conditional on age and age-20 AFQT. In other words, $\beta_3$ is not a between-wave shift per se; it is the PE for the second assessment at wave 4.

#### *Use of coefficients for practice effect adjustment*

An important point is that the practice effect estimates $\beta_3$ through $\beta_8$ are not additive. The combination of wave and number of prior assessments are captured by individual terms in the model. For example, the expected practice effect for an individual taking a test for the third time at wave 3 would correspond to $\beta_6$ (additionally adjusting with $\beta_{s1}$ and/or $\beta_{s2}$ if any prior assessments were skipped). It would *not* be calculated taking the cumulative effect of practice effects at wave 2 and 3 (i.e., $\beta_4 + \beta_6$).

An additional clarification is that "skipped" assessments are only counted for individuals that missed a wave *after* enrollment into the study. Thus, waves occurring prior to an attrition replacement's entry into the study are not counted as skipped.

***Cognitive factor score composites***

Performance across 5 cognitive domains was assessed using factor scores based on previous latent variable analyses of multiple neuropsychological tests. Higher scores reflect better performance in each domain. Details of each factor model are described in previous publications [1-4], and brief summaries are provided below:

*Episodic memory:* The episodic memory factor score [4] was based on the California Verbal Learning Test-II [CVLT-II; 5] short delay free recall, long delay free recall, and the total score for the learning trials, as well as Wechsler Memory Scale-III Logical Memory and WMS-III Visual Reproductions subtests [6] immediate and long delay recall.

*Executive function:* The executive function factor score [2, 7] was based on the Stroop interference test [8], letter-number sequencing and digit span subtests of the Wechsler Memory Scale (WMS)-III [9], the Reading Span Task [10], Trail Making Test Switching task and the Category Switching trial from Delis-Kaplan Executive Function System (D-KEFS) [11]. Inhibition and shifting scores were adjusted for performance on the baseline conditions.

*Verbal fluency:* The verbal fluency factor score [1] was based on Letter (F, A, S) and Category (animals, boys' names, fruits/furniture) fluency trials from the D-KEFS [11]

*Processing Speed:* The processing speed factor score [3] was based on the number sequencing and letter sequencing conditions from the Delis-Kaplan Executive Function System [D-KEFS; 11], word reading and color naming conditions from the Stroop task [8], and simple and choice reaction time tasks.

*Visuospatial ability:* The visuospatial factor score was based on the Gottschaldt Hidden Figures task [12], the Mental Rotation task [13], and the Box-Folding component of the Armed Forces Qualifying Test [14] that was given at each VETSA assessment (see below).

***Armed Forces Qualification Test (AFQT)***

General cognitive ability (GCA) was measured with the validated Armed Forces Qualification Test (AFQT) [15]. The AFQT is a 100-item multiple-choice paper-and-pencil test administered during military induction at average age 20. The AFQT is highly correlated with standard IQ measures (r=.84) [16]. AFQT scores from military induction (hereafter referred to as "age 20 AFQT") were reported as percentile scores and have been transformed to the standard normal distribution using a probit transform.

The AFQT was also assessed at each VETSA assessment (the Box-Folding component was used for the visuospatial factor score composite). To reduce burden on participants, only half of the items were given at waves 3 and 4. Therefore, we used the "half-scale" scores from all waves for consistency. The correlation between half-scale and full-scale scores was 0.95 and 0.96, respectively.

***Classification of mild cognitive impairment***

Mild cognitive impairment (MCI) was diagnosed using the Jak-Bondi approach based on up to 19 neuropsychological tests covering 6 cognitive abilities (see **Supplemental Table S4** for assignment of scores to abilities) [17, 18]. Note that the domains and tests that they comprise are different than those used in the cognitive factor score composites described above. For purposes of MCI diagnosis, composite scores for certain tests were calculated by averaging individual component scores (i.e., immediate and delayed recall from) to reduce the imbalance in number of tests per ability. Each test was administered at all waves with two exceptions: spatial span was administered only at waves 1-3, and Boston Naming Test was administered only at waves 3 and 4. The impairment criterion was scoring>1.5 SDs below publisher provided age-adjusted normative means on 2 or more tasks within a cognitive ability. This threshold is stricter than the more commonly used threshold of >1 SD as we have shown this provides a more reasonable prevalence in in the age range of our  community dwelling sample [19], and we have shown this MCI classification is related to Alzheimer's disease genetic risk [20] and an Alzheimer's-related brain structure [21].

### SUPPLEMENTAL TABLES AND FIGURES

**Supplemental Table S1. Interpretation of GEE model coefficients.**

| Term | Interpretation |
|---|---|
| **$\beta_{age}$, $\beta_{afqt}$** | The effect of a one unit increase in age or age 20 AFQT score on cognitive outcome, respectively. The coefficients below can therefore be interpreted as being conditional on age and early life general cognitive ability. |
| **$\beta_0$** | The reference (intercept) will be wave 1 and assessment 1. |
| **$\beta_{s1}$, $\beta_{s2}$** | Effect of skipping one or two waves immediately prior to the current assessment, respectively. |
| **$\beta_1$** | This can be interpreted as the difference between wave 2 replacement subjects' baseline score and the original samples' baseline score. Although coded as the main effect of wave 2, due to inclusion of interaction terms for assessment and wave, this coefficient corresponds to the reference level of assessment at wave 2. In the case of wave 2, that corresponds to the effect of the first assessment at wave 2. |
| **$\beta_2$** | This can be interpreted as the difference between wave 3 replacement subjects' baseline score and the original samples' baseline score. Although coded as the main effect of wave 3, due to inclusion of interaction terms for assessment and wave, this coefficient corresponds to the reference level of assessment at wave 3. In the case of wave 3, that corresponds to the effect of the first assessment at wave 3. |
| **$\beta_3$** | This can be interpreted as the practice effect for assessment 2 occurring at wave 4. Although coded as the main effect of wave 4, due to inclusion of interaction terms for assessment and wave, this coefficient corresponds to the reference level of assessment at wave 4. Note, this coefficient is interpreted differently than $\beta_1$ and $\beta_2$ because no participants had their first assessment at wave 4. Adding a separate term for [*w*=4 & *a*=2] would induce collinearity, so was omitted from the model. Thus, the reference level corresponds to the *second* assessment occurring at wave 4 |
| **$\beta_4$** | The practice effect occurring at wave 2, 2$^{nd}$ assessment. |
| **$\beta_5$** | The practice effect occurring at wave 3, 2$^{nd}$ assessment. |
| **$\beta_6$** | The practice effect occurring at wave 3, 3$^{rd}$ assessment. |
| **$\beta_7$** | The practice effect occurring at wave 4, 3$^{rd}$ assessment. |
| **$\beta_8$** | The practice effect occurring at wave 4, 4$^{th}$ assessment. |

**Supplemental Table S2. Unstandardized practice effect estimates.** Practice effect estimates are presented on unstandardized raw score scale. Models were fit using all participants and assessments patterns but only estimates corresponding to practice effect for the group of individuals who attended all four waves are presented here (i.e., column 1 = $\beta_4$, column 2 = $\beta_6$, column 3 = $\beta_8$).

| | Follow-up 1 | | | Follow-up 2 | | | Follow-up 3 | | |
|---|---|---|---|---|---|---|---|---|---|
| **Measure** | **Estimate** | **SE** | **95% CI** | **Estimate** | **SE** | **95% CI** | **Estimate** | **SE** | **95% CI** |
| CVLT Long Delay | 0.346 | 0.211 | [-0.07, 0.76] | 0.555 | 0.283 | [ 0.00, 1.11] | 0.212 | 0.310 | [-0.39, 0.82] |
| CVLT Short Delay | 0.428 | 0.219 | [ 0.00, 0.86] | 0.467 | 0.266 | [-0.05, 0.99] | 0.654 | 0.332 | [ 0.00, 1.30] |
| CVLT Trials 1-5 | 1.851 | 0.638 | [ 0.60, 3.10] | 1.526 | 0.840 | [-0.12, 3.17] | 0.750 | 0.943 | [-1.10, 2.60] |
| Logical Mem Delay | 1.571 | 0.486 | [ 0.62, 2.52] | 1.091 | 0.609 | [-0.10, 2.29] | 2.317 | 0.673 | [ 1.00, 3.64] |
| Logical Mem Immediate | 1.625 | 0.432 | [ 0.78, 2.47] | 0.879 | 0.560 | [-0.22, 1.98] | 1.577 | 0.620 | [ 0.36, 2.79] |
| Vis Repro Delay | 3.983 | 1.326 | [ 1.38, 6.58] | 5.402 | 1.613 | [ 2.24, 8.56] | 2.853 | 2.001 | [-1.07, 6.77] |
| Vis Repro Immediate | 1.172 | 0.883 | [-0.56, 2.90] | 4.190 | 1.127 | [ 1.98, 6.40] | 4.399 | 1.495 | [ 1.47, 7.33] |
| Cat. Switching (total correct) | 0.320 | 0.266 | [-0.20, 0.84] | -0.117 | 0.277 | [-0.66, 0.43] | 0.205 | 0.329 | [-0.44, 0.85] |
| Digit Span Total | 0.623 | 0.230 | [ 0.17, 1.08] | -0.212 | 0.279 | [-0.76, 0.33] | 0.121 | 0.342 | [-0.55, 0.79] |
| Letter-Num Sequencing | 0.159 | 0.162 | [-0.16, 0.48] | 0.224 | 0.215 | [-0.20, 0.65] | -0.107 | 0.236 | [-0.57, 0.36] |
| Reading Span | 0.348 | 0.334 | [-0.31, 1.00] | 0.620 | 0.397 | [-0.16, 1.40] | 0.646 | 0.516 | [-0.37, 1.66] |
| Stroop Interference | -0.179 | 0.565 | [-1.29, 0.93] | 1.108 | 0.741 | [-0.34, 2.56] | 1.153 | 0.686 | [-0.19, 2.50] |
| Trails 4 (time) | -0.067 | 0.028 | [-0.12, -0.01] | -0.052 | 0.035 | [-0.12, 0.02] | -0.016 | 0.048 | [-0.11, 0.08] |
| Cat. Switching (# switches) | 0.151 | 0.215 | [-0.27, 0.57] | 0.062 | 0.223 | [-0.37, 0.50] | 0.110 | 0.264 | [-0.41, 0.63] |
| Category Fluency | 0.229 | 0.575 | [-0.90, 1.36] | -0.050 | 0.720 | [-1.46, 1.36] | 0.878 | 0.781 | [-0.65, 2.41] |
| Letter Fluency | 0.134 | 0.754 | [-1.34, 1.61] | 0.385 | 0.994 | [-1.56, 2.33] | -0.227 | 1.057 | [-2.30, 1.84] |
| Choice RT (time) | 0.009 | 0.007 | [ 0.00, 0.02] | -0.014 | 0.010 | [-0.03, 0.01] | 0.009 | 0.014 | [-0.02, 0.04] |
| Simple RT (time) | 0.008 | 0.008 | [-0.01, 0.02] | -0.033 | 0.009 | [-0.05, -0.02] | 0.008 | 0.016 | [-0.02, 0.04] |
| Stroop Color | 1.590 | 0.669 | [ 0.28, 2.90] | 1.813 | 0.958 | [-0.06, 3.69] | 1.395 | 1.031 | [-0.63, 3.42] |
| Stroop Word | 1.205 | 0.894 | [-0.55, 2.96] | -0.097 | 1.202 | [-2.45, 2.26] | -1.061 | 1.432 | [-3.87, 1.74] |
| Trails 2 (time) | -0.032 | 0.025 | [-0.08, 0.02] | -0.037 | 0.029 | [-0.09, 0.02] | -0.051 | 0.038 | [-0.13, 0.02] |
| Trails 3 (time) | -0.026 | 0.025 | [-0.07, 0.02] | -0.035 | 0.035 | [-0.10, 0.03] | -0.018 | 0.044 | [-0.10, 0.07] |
| AFQT Box Folding | 0.222 | 0.060 | [ 0.10, 0.34] | 0.164 | 0.074 | [ 0.02, 0.31] | 0.023 | 0.084 | [-0.14, 0.19] |
| Hidden Figures | 1.918 | 0.486 | [ 0.96, 2.87] | 2.123 | 0.573 | [ 1.00, 3.25] | 2.644 | 0.541 | [ 1.58, 3.70] |
| Mental Rotation | 3.884 | 0.903 | [ 2.11, 5.65] | 3.890 | 1.092 | [ 1.75, 6.03] | 3.036 | 1.360 | [ 0.37, 5.70] |
| Boston Naming Test* | 0.632 | 0.231 | [ 0.18, 1.09] | - | - | - | - | - | - |
| Matrix Reasoning | 0.394 | 0.395 | [-0.38, 1.17] | 1.325 | 0.534 | [ 0.28, 2.37] | 1.575 | 0.620 | [ 0.36, 2.79] |
| Spatial Span† | 0.761 | 0.202 | [ 0.36, 1.16] | -0.112 | 0.273 | [-0.65, 0.42] | - | - | - |
| Trails 1 (time) | 0.004 | 0.016 | [-0.03, 0.04] | 0.026 | 0.022 | [-0.02, 0.07] | -0.025 | 0.029 | [-0.08, 0.03] |
| Vis Repro Copy | 0.317 | 0.342 | [-0.35, 0.99] | -0.016 | 0.406 | [-0.81, 0.78] | 1.409 | 0.617 | [ 0.20, 2.62] |

* Boston Naming Test was administered at waves 3 and 4 only. The estimate for follow-up 1 therefore corresponds to $\beta_3$.

† Spatial Span was administered at waves 1, 2, and 3 only.

**Supplemental Table S3. Effect of practice effect adjustment on cognitive composite scores at follow-up.** Paired t-tests were used to examine the difference in cognitive composite scores calculated from practice effect-adjusted and unadjusted measures. All scores were standardized using the sample means and standard deviations at wave 1. Therefore, the mean difference can be interpreted as the average difference after practice effect-adjustment in wave 1 standard deviation units.

| Domain | Wave | t | df | p | Mean Difference | 2.5% CI | 97.5% CI |
|---|---|---|---|---|---|---|---|
| Episodic memory | 2 | 77.131 | 1,202.000 | <0.001 | 0.214 | 0.209 | 0.220 |
| | 3 | 76.008 | 1,170.000 | <0.001 | 0.218 | 0.212 | 0.223 |
| | 4 | 115.769 | 835.000 | <0.001 | 0.251 | 0.247 | 0.255 |
| Executive function | 2 | 90.634 | 1,202.000 | <0.001 | 0.159 | 0.156 | 0.163 |
| | 3 | 89.442 | 1,169.000 | <0.001 | 0.144 | 0.141 | 0.148 |
| | 4 | 55.630 | 835.000 | <0.001 | 0.162 | 0.156 | 0.168 |
| Fluency | 2 | 79.284 | 1,202.000 | <0.001 | 0.048 | 0.047 | 0.050 |
| | 3 | 73.064 | 1,170.000 | <0.001 | 0.035 | 0.034 | 0.036 |
| | 4 | 26.513 | 835.000 | <0.001 | 0.073 | 0.067 | 0.078 |
| Processing speed | 2 | 57.786 | 1,200.000 | <0.001 | 0.088 | 0.085 | 0.091 |
| | 3 | 61.609 | 1,165.000 | <0.001 | 0.152 | 0.147 | 0.157 |
| | 4 | 21.345 | 832.000 | <0.001 | 0.101 | 0.091 | 0.110 |
| Visuospatial | 2 | 77.283 | 1,199.000 | <0.001 | 0.223 | 0.218 | 0.229 |
| | 3 | 62.690 | 1,164.000 | <0.001 | 0.193 | 0.187 | 0.199 |
| | 4 | 77.709 | 831.000 | <0.001 | 0.204 | 0.199 | 0.209 |

**Supplemental Table S4. Neuropsychological tests and scores used to define mild cognitive impairment.**

| Cognitive Domain | Tests and Measures | No. of Measures |
|---|---|---|
| Episodic Memory | CVLT-II: Sum of trials 1-5; delayed free recall (composite)*<br>WMS-III: Logical memories immediate, delayed free recall (composite)*<br>WMS-III: Visual reproduction immediate and delayed free recall (composite)* | 3 |
| Executive Function | DKEFS Trails: Switching (condition 4)<br>DKEFS Fluency: Category switching<br>Stroop: Color-word, interference<br>WASI: Matrix reasoning | 4 |
| Attention/ Working Memory | WMS-III: Digit span<br>WMS-III: Spatial span†<br>WMS-III: Letter-number sequencing<br>DKEFS Trails: Cancellations (condition 1) | 4 |
| Verbal/Language | DKEFS: Letter fluency<br>DKEFS: Category fluency<br>Boston Naming Test‡ | 3 |
| Visuospatial | Gottschaldt Hidden Figures<br>Card rotation<br>WMS-III: Visual reproduction copy | 3 |
| Processing Speed | DKEFS Trails: Number sequencing, letter sequencing (conditions 2 and 3 composite)*<br>Stroop: Word condition, color condition (composite)* | 2 |

Note: CVLT-II, California Verbal Learning Test –Version II; WMS-III, Wechsler Memory Scale–Version III; DKEFS, Delis-Kaplan Executive Function System; WASI, Wechsler Abbreviated Scale of Intelligence.

* Composite scores for tests were calculated by taking the average of individual component scores (e.g., immediate and delayed recall portions of the test).

† Spatial span was administered at waves 1, 2, and 3.

‡ Boston Naming Test was administered at waves 3 and 4.

**Figure 1. Summary of assessments and age distributions by wave.** A) Upset plot describing the patterns of baseline and follow-up assessments completed. Individuals with baseline assessments at waves 2 and 3 were recruited as attrition replacements. B) Density plots of age distributions at each wave. Attrition replacement (AR) participants were recruited at waves 2 and 3 and were age-matched to the on-going longitudinal sample at the corresponding timepoint.

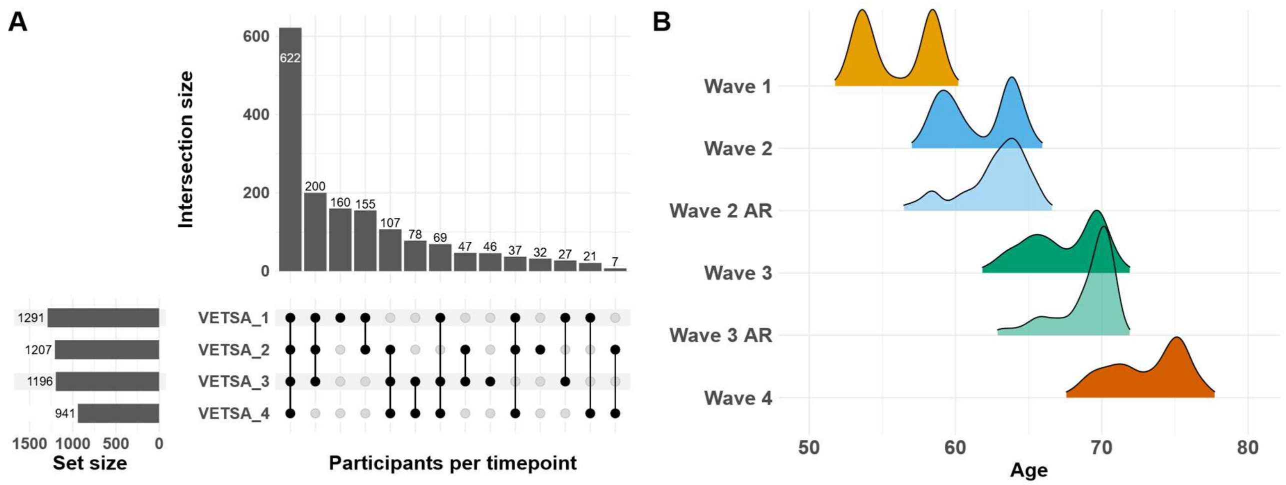
